# Latent Classes of Cannabis, ENDS, and Tobacco Use and Environmental Exposure Among Adolescents and Young Adults in the ECHO Cohort

**DOI:** 10.64898/2026.04.03.26350120

**Authors:** Hannah J. Appleseth, John M. Felt, Amy M. Cohn, Rebecca J. Schmidt, Julie M. Croff, Thad R. Leffingwell

## Abstract

**Purpose:** Polyuse of tobacco, nicotine, and cannabis products has increased among adolescents and young adults, but less is known about the prevalence of environmental (i.e., secondhand) exposure to these products. This study aimed to identify groups of tobacco, electronic nicotine delivery systems, and cannabis use and environmental exposures among adolescents and young adults.

**Methods:** Data were collected between 2018 and 2022 from adolescents and young adults enrolled in the Environmental influences on Child Health Outcomes (ECHO) research program. A multiple-group latent class analysis characterized age group (early, middle, or late adolescence/young adulthood) patterns of environmental exposure to cannabis smoke, tobacco smoke, and electronic nicotine delivery system aerosol emissions (≥1 hour/day or cohabitation with daily indoor use), and personal use of cannabis, tobacco, and electronic nicotine delivery systems, categorized as experimental (<1 time/month) or current (≥1 time/month).

**Results:** Among 2,876 participants from 19 ECHO cohorts (*M_age_*= 17.28 years; 52% [*n*=1,495] female, 9.1% [*n*=248] Hispanic, 58.4% [n=1,583] non-Hispanic White), four classes were identified: No Use/No Exposure (53%), No Use/Polyexposure (10%), Experimental Use/Low Exposure (22%), and Polysubstance Use/Polyexposure (14%). The No Use/No Exposure and No Use/Polyexposure classes primarily included early and middle adolescents, whereas the Experimental Use/Low Exposure and Polysubstance Use/Polyexposure classes primarily included late adolescents and young adults.

**Conclusions:** Our findings suggest that investigating environmental exposures alongside polyuse is an emerging priority for tobacco and cannabis research and prevention. Prospective data from future ECHO releases can extend our findings by characterizing polyuse and polyexposure over time and identifying correlates of risk.

---

It took nearly a century to fully realize and address the health risks of cigarettes and environmental tobacco smoke exposure (ETSE).^1^ Fortunately, smoke-free air policies and messaging have contributed to increased smoking cessation and decreased ETSE over the past three decades.^2^ Despite these advances, over one-third of adolescents and young adults still experience ETSE, and 10% of US young adults (18-24 years) and 1.2% of adolescents (12-17 years) report past month cigarette use.^3^

New concerns have emerged due to the rapid expansion of cannabis legalization and the proliferation of electronic nicotine delivery systems (ENDS).^4^ Six percent of adolescents and 25% of young adults reported past month cannabis use in 2024, and smoking and vaping are the most common modalities.^5,6^ Similarly, 6% of adolescents and approximately 25% of young adults reported past month ENDS use, and 40-50% of adolescents and young adults who use ENDS also use cannabis.^5^ ENDS and cannabis use have been independently linked to increased odds of cigarette smoking initiation;^7,8^ 15.5% of US young adults reported past 30-day co-use of cigarettes and cannabis, and among those who used ENDS, 31.9% reported past 30-day co-use with other tobacco products.^9^

The prevalence of environmental cannabis smoke exposure (ECSE) and environmental nicotine aerosol exposure (ENAE) appear to have also increased. Past 30-day ENAE among U.S. adolescents rose from 25.2% in 2015 to 33.2% in 2018.^10^ Comparable estimates for ECSE among adolescents are limited, as most studies have focused on small, high-risk samples of children,^11^ however, a recent study found that nearly half of young adults surveyed reported past 30-day ECSE.^12^

Perhaps the largest changes have been observed among adults in the age groups most likely to be parents of adolescents. Among US adults ages 35-49, past 30-day cannabis use more than doubled from 7.7% in 2014 to 18.9% in 2024, while past 30-day cigarette use declined from 25.0% to 18.5%.^5^ Similarly, among adults ages 50-64, past 30-day cannabis use more than doubled from 5.9% in 2014 to 13.6% in 2024, while cigarette use declined from 22.3% to 17.7%.^5^ Although the National Survey of Drug Use and Health (NSDUH) only began tracking ENDS use in 2022, recent data indicate that in 2024, 11.2% of adults ages 35-49 and 3.8% of adults ages 50-64 reported past month ENDS use.^5^ Furthermore, adults who smoke cigarettes are more likely to initiate and sustain cannabis and tobacco poly-use patterns.^9^

Shifts in adult tobacco and cannabis use patterns could have downstream implications for youth. Cannabis and ENDS emissions carry many of the same respiratory irritants, fine particles, volatile organic compounds (VOCs), and toxicants as tobacco smoke, and exposure to cannabis and ENDS emissions have been linked to increased respiratory symptoms and asthma exacerbation.^12,13^ Moreover, early evidence shows that polyexposure to these products can compound the respiratory and cardiovascular risks associated with each substance individually, including oxidative stress, endothelial dysfunction, and pulmonary inflammation.^14^

Given these trends, we need to identify and understand distinct profiles of polyuse and polyexposure, particularly among adolescents and young adults. Substance use is most likely to be initiated during adolescence, and adolescents and young adults could potentially experience multiple exposures from caregivers in the home and peers in social environments.^15^ The use of a person-centered analytic approach to examine how these patterns vary by adolescent stage will allow for an improved understanding of the scope of the problem and inform age-appropriate intervention strategies.

## The current study

We conducted secondary analyses using data from 19 U.S.-based pediatric cohorts enrolled in the Environmental influences on Child Health Outcomes (ECHO) study to complete the following aims: (1) Characterize underlying use and environmental exposure patterns to tobacco, ENDS, and cannabis among adolescents and young adults, and (2) Compare cross-sectional substance use and environmental exposure latent classes across adolescent stages, as defined by the National Academies of Sciences, which includes: early (ages 11-13 years), middle (ages 14-17 years), and late adolescence/young adulthood (ages 18-24 years).^16,17^

## METHODS

### Participants

The sample (*N*=2,876) included adolescents and young adults (ages 11-24) enrolled in the Environmental influences on Child Health Outcomes (ECHO) program.^18^ Figure 1 details participant inclusion, exclusion, and deduplication procedures. Participants were retained if they completed the standardized ECHO-Wide Cohort Data Collection Protocol (EWCP) to promote measurement consistency across study sites and data collection years.^16^ Supplementary Tables S1-S3 show substance use and environmental exposures stratified by survey year (Table S1), year of birth (Table S2), and cohort (Table S3).

**Figure 1.**
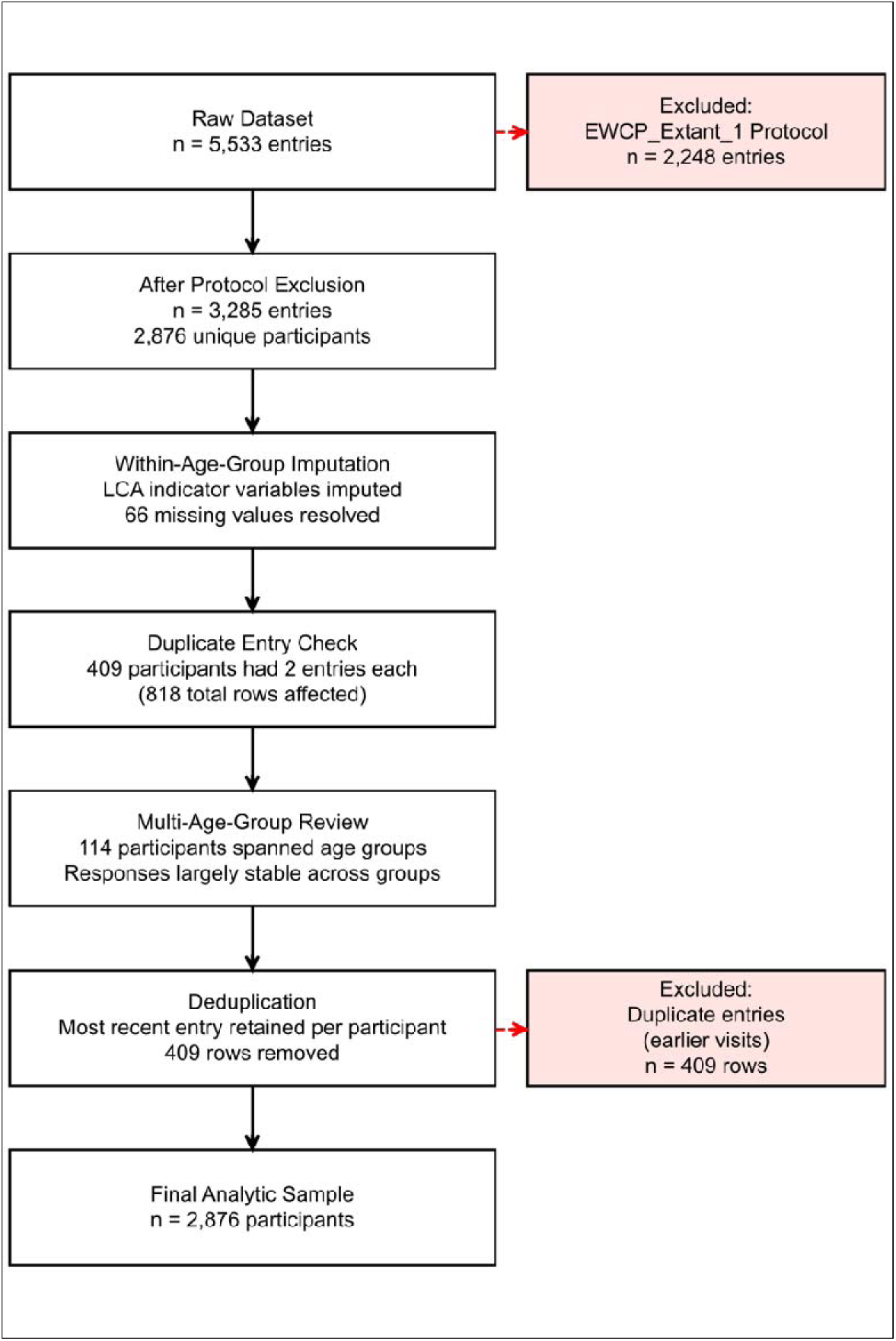
Participant Flow Diagram.

### Procedures

Adult participants (18+ years) provided written consent. Primary caregivers provided consent for participants aged 11-17 years, and children provided assent. Per our data use agreement, potential participant and cohort geographical data were not available for the current analyses. Each study site received local institutional review board (IRB) approval, and the ECHO-wide protocol was approved by a single IRB. Additionally, this study was approved by the [university name redacted] Institutional Review Board (#XXXXX).

### Measures

#### Sociodemographic characteristics

Participants reported their age, gender, date of birth, race (American Indian or Alaska Native, Asian, Black or African American, Native Hawaiian or Pacific Islander, White, or more than one race), ethnicity (Hispanic or Latino, Non-Hispanic or Latino), maternal education level (less than a high school diploma or GED, high school diploma or GED, some college but no degree/associate degree/trade school, bachelor’s degree, graduate degree), and housing type (single-family/detached home; duplex/triplex; row house; low-rise apartment [1-3 floors]; high-rise apartment [3+ floors]; mobile home/trailer; other).

Race and ethnicity were collapsed to 4 categories including: Hispanic (any race), non-Hispanic White, non-Hispanic Black, and non-Hispanic other race or multiracial. Housing type was collapsed into 3 categories: single-family/detached home; multi-unit/apartment (duplex/triplex, low-rise apartment, high-rise apartment); and other (row house, mobile home/trailer, other).

#### Tobacco, ENDS, and cannabis use and environmental exposures

The Youth Risk Behavior Survey-Substance Use module was used to collect substance use and environmental exposure data.^19^ Lifetime use of cigarettes, ENDS, and cannabis were assessed via binary items (yes/no; “Have you ever tried cigarettes?”; “Have you ever used an electronic vapor product such as ENDS, e-cigars, or e-pipes?”; “Have you ever tried marijuana, including hashish? Include smoking, vaping, or eating.”). Participants who reported lifetime use were then asked to report their substance use over the past 12 months using a 6-point scale (1 = *never*; 2 = *less than once a month*; 3 = *less than once a week*; 4 = *1-2 days per week*; 5 = *3-5 days per week*; 6 = *almost every day/every day*).

Cigarettes and other tobacco products were combined into a single variable that reflected the maximum reported frequency across products. Because of the low prevalence of tobacco use, it was dichotomized (0 = *never*; 1 = *any lifetime use*) for the latent class analysis to reduce sparsity. ENDS and cannabis use were categorized as 0 = *never*; 1 = *experimental use* (lifetime use but not within the past month); or 2 = *current use* (daily, weekly, or past month use).

Environmental tobacco smoke exposure (ETSE), environmental nicotine aerosol exposure (ENAE), and environmental cannabis smoke exposure (ECSE) were assessed by: (1) average daily hours spent around others smoking or vaping (“How many hours per day, on average, are you around someone who is smoking [cigarettes/ENDS/marijuana], close enough to smell the smoke or vapor?”) and/or (2) the number of household members who regularly smoke or vape indoors (“How many other people, not counting yourself, who live in your home regularly smoke [cigarettes/ENDS/marijuana] inside the house?”). Responses were dichotomized as 0 = *no exposure* (0 hours per day and 0 household members who smoke or vape indoors) or 1 = *exposure* (≥1 hours per day and/or ≥1 household member who smokes or vapes indoors).

### Statistical analyses

Analyses were conducted in RStudio, version 2025.05.1+513 using the mglca, poLCA, ggplot2, and dplyr packages.^20,21^

#### Descriptives

Frequencies, percentages, chi-squares, and Cramér’s V were calculated to assess prevalences and differences across age groups. To assess combinations of substance, use and environmental exposures, two mutually exclusive eight-category variables were constructed reflecting any past use (none; each product alone; and each dual and triple combination of tobacco, ENDS, and cannabis; *N* = 2,773) and any environmental exposure (none; each product alone; and each dual and triple combinations of ETSE, ENAE, and ECSE; *N* = 2,619) and then compared across age groups. Post-hoc analyses identified which categories differed across age groups and the Holm-Bonferroni procedure was used to control familywise error.

#### Participant Selection and Missing Data

Details of participant inclusion, exclusion, and deduplication are described in Figure 1. Missingness across indicators was <10%. Logistic regression models confirmed that missingness was significantly associated with race/ethnicity and maternal education, consistent with a Missing at Random (MAR) assumption. Participants with partial data were retained in the latent class analysis via full-information maximum likelihood estimation.^22^

#### Multiple-Group Latent Class Analyses

A multiple-group LCA stratified by age group (early adolescence [11-13 years], middle adolescence [14-17 years], and late adolescence/young adulthood [18-24 years]) was performed using six indicators: tobacco use, cannabis use, ENDS use, ETSE, ENAE, and ECSE. A 4-class invariant model demonstrated the best fit and parsimony among models specifying 2-6 classes based on the Bayesian Information Criterion (BIC), Akaike Information Criterion (AIC), log-likelihood, and entropy metrics (Supplementary Table S4; Supplementary Table S5). Modal assignment was used to assign class membership and average posterior probabilities of correct assignment ranging from 0.81 to 0.92 across classes.

## RESULTS

### Sample characteristics

Table 1 presents sample sociodemographic characteristics stratified by age group, including early adolescents (ages 11-13; *n*=226, 7.9%), middle adolescents (ages 14-17; *n*=1,248, 43.4%), and late adolescents/young adults (ages 18-24; *n*=1,402, 48.7%). The sample was 9.1% (*n*=248) Hispanic, 58.4% (*n*=1583) non-Hispanic White, and 52% (*n*=1495) female. Over two-thirds resided in a single-family home (66.7%, *n*=1,246) and 47.3% (*n*=1,034) reported their mother’s highest education level was a bachelor’s degree or more.

**Table 1.**
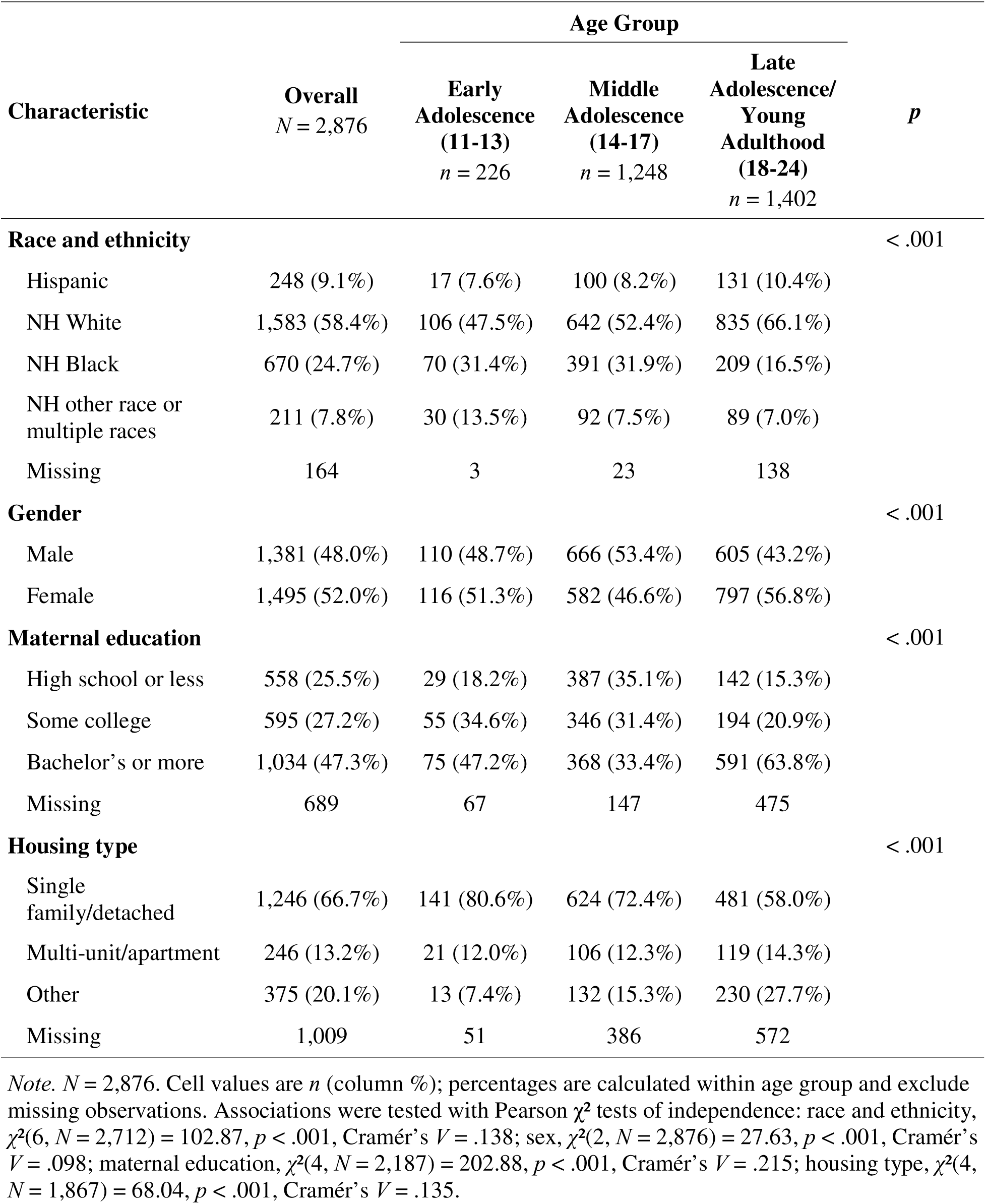
Sample Characteristics by Age Group.

| Characteristic | Overall<br>*N* = 2,876 | Age Group |  |  | *p* |
| --- | --- | --- | --- | --- | --- |
|  |  | Early<br>Adolescence<br>(11-13)<br>*n* = 226 | Middle<br>Adolescence<br>(14-17)<br>*n* = 1,248 | Late<br>Adolescence/<br>Young<br>Adulthood<br>(18-24)<br>*n* = 1,402 |  |
| **Race and ethnicity** |  |  |  |  | < .001 |
| Hispanic | 248 (9.1%) | 17 (7.6%) | 100 (8.2%) | 131 (10.4%) |  |
| NH White | 1,583 (58.4%) | 106 (47.5%) | 642 (52.4%) | 835 (66.1%) |  |
| NH Black | 670 (24.7%) | 70 (31.4%) | 391 (31.9%) | 209 (16.5%) |  |
| NH other race or<br>multiple races | 211 (7.8%) | 30 (13.5%) | 92 (7.5%) | 89 (7.0%) |  |
| Missing | 164 | 3 | 23 | 138 |  |
| **Gender** |  |  |  |  | < .001 |
| Male | 1,381 (48.0%) | 110 (48.7%) | 666 (53.4%) | 605 (43.2%) |  |
| Female | 1,495 (52.0%) | 116 (51.3%) | 582 (46.6%) | 797 (56.8%) |  |
| **Maternal education** |  |  |  |  | < .001 |
| High school or less | 558 (25.5%) | 29 (18.2%) | 387 (35.1%) | 142 (15.3%) |  |
| Some college | 595 (27.2%) | 55 (34.6%) | 346 (31.4%) | 194 (20.9%) |  |
| Bachelor's or more | 1,034 (47.3%) | 75 (47.2%) | 368 (33.4%) | 591 (63.8%) |  |
| Missing | 689 | 67 | 147 | 475 |  |
| **Housing type** |  |  |  |  | < .001 |
| Single<br>family/detached | 1,246 (66.7%) | 141 (80.6%) | 624 (72.4%) | 481 (58.0%) |  |
| Multi-unit/apartment | 246 (13.2%) | 21 (12.0%) | 106 (12.3%) | 119 (14.3%) |  |
| Other | 375 (20.1%) | 13 (7.4%) | 132 (15.3%) | 230 (27.7%) |  |
| Missing | 1,009 | 51 | 386 | 572 |  |

*Note.* *N* = 2,876. Cell values are *n* (column %); percentages are calculated within age group and exclude missing observations. Associations were tested with Pearson $\chi^2$ tests of independence: race and ethnicity, $\chi^2(6, N = 2,712) = 102.87, p < .001$ , Cramér's *V* = .138; sex, $\chi^2(2, N = 2,876) = 27.63, p < .001$ , Cramér's *V* = .098; maternal education, $\chi^2(4, N = 2,187) = 202.88, p < .001$ , Cramér's *V* = .215; housing type, $\chi^2(4, N = 1,867) = 68.04, p < .001$ , Cramér's *V* = .135.

### Frequency of substance use and environmental exposures

Table 2 presents substance use and environmental exposure frequencies by age group. Cannabis was the most used substance (19% experimental, *n* = 530; 15% current, *n* = 436), followed by ENDS (16% experimental, *n* = 454; 10% current, *n* = 282), and then tobacco (12% experimental, *n* = 339; 3.4% current, *n* = 97). Current use of all three substances increased with age, with the largest increase for cannabis, which rose from 0.5% (*n* = 1) in early adolescence to 8.6% (*n* = 106) in middle adolescence, and 24% (*n* = 329) in late adolescence/young adulthood. Across the same age groups, current ENDS use rose from 1.4% (*n* = 3) to 8.2% (*n* = 101) to 13% (*n* = 178), and current tobacco use from 1.3% (*n* = 3) to 2.9% (*n* = 36) to 4.2% (*n* = 58).

**Table 2.** Substance Use Frequency and Exposure by Age Group.

|  | **Overall**<br>**N = 2876<sup>1</sup>** | **Early**<br>**Adolescence**<br>**N = 226<sup>1</sup>** | **Middle**<br>**Adolescence**<br>**N = 1248<sup>1</sup>** | **Late**<br>**Adolescence/Young**<br>**Adult**<br>**N = 1402<sup>1</sup>** | **p-**<br>**value<sup>2</sup>** |
| --- | --- | --- | --- | --- | --- |
| **ENDS frequency** |  |  |  |  | <0.001 |
| Never | 2,047 (74%) | 207 (94%) | 972 (79%) | 868 (65%) |  |
| Experimental | 454 (16%) | 11 (5.0%) | 160 (13%) | 283 (21%) |  |
| Current Use | 282 (10%) | 3 (1.4%) | 101 (8.2%) | 178 (13%) |  |
| **Cannabis frequency** |  |  |  |  | <0.001 |
| Never | 1,852 (66%) | 209 (95%) | 979 (80%) | 664 (49%) |  |
| Experimental | 530 (19%) | 9 (4.1%) | 145 (12%) | 376 (27%) |  |
| Current Use | 436 (15%) | 1 (0.5%) | 106 (8.6%) | 329 (24%) |  |
| **Tobacco frequency** |  |  |  |  | <0.001 |
| Never | 2,397 (85%) | 215 (95%) | 1,106 (89%) | 1,076 (79%) |  |
| Experimental | 339 (12%) | 8 (3.5%) | 95 (7.7%) | 236 (17%) |  |
| Current Use | 97 (3.4%) | 3 (1.3%) | 36 (2.9%) | 58 (4.2%) |  |
| **Cigarette frequency** |  |  |  |  | <0.001 |
| Never | 2,462 (87%) | 214 (96%) | 1,143 (93%) | 1,105 (81%) |  |
| Experimental | 309 (11%) | 7 (3.1%) | 78 (6.3%) | 224 (16%) |  |
| Current Use | 54 (1.9%) | 2 (0.9%) | 12 (1.0%) | 40 (2.9%) |  |
| **Smokeless tobacco**<br>**frequency** |  |  |  |  | 0.229 |
| Never | 2,108 (97%) | 214 (99%) | 1,180 (98%) | 714 (96%) |  |
| Experimental | 33 (1.5%) | 1 (0.5%) | 16 (1.3%) | 16 (2.2%) |  |
| Current Use | 22 (1.0%) | 1 (0.5%) | 11 (0.9%) | 10 (1.4%) |  |
| **Cigar / Cigarillo /**<br>**Little cigar frequency** |  |  |  |  | 0.038 |
| Never | 2,065 (96%) | 212 (99%) | 1,160 (96%) | 693 (94%) |  |
| Experimental | 60 (2.8%) | 1 (0.5%) | 29 (2.4%) | 30 (4.1%) |  |
| Current Use | 32 (1.5%) | 2 (0.9%) | 18 (1.5%) | 12 (1.6%) |  |
| **Environmental Exposures** |  |  |  |  |  |
| **ETSE** |  |  |  |  | <0.001 |
| No Exposure | 2,157 (81%) | 161 (75%) | 892 (77%) | 1,104 (86%) |  |
| Exposure | 506 (19%) | 54 (25%) | 267 (23%) | 185 (14%) |  |
| **ENAE** |  |  |  |  | <0.001 |
| No Exposure | 2,162 (81%) | 177 (82%) | 977 (84%) | 1,008 (78%) |  |
| Exposure | 504 (19%) | 38 (18%) | 181 (16%) | 285 (22%) |  |
| **ECSE** |  |  |  |  | <0.001 |
| No Exposure | 2,138 (80%) | 179 (86%) | 1,005 (87%) | 954 (74%) |  |
| Exposure | 526 (20%) | 28 (14%) | 156 (13%) | 342 (26%) |  |

*Note.* ENDS= Electronic nicotine delivery system, ETSE=Environmental tobacco smoke exposure, ENAE=Environmental nicotine aerosols exposure, ECSE=Environmental cannabis smoke exposure. Experimental use=any lifetime use or past 12-month use but no past-month use; Current use=daily, weekly, or past month use.
<sup>1</sup>n (%) Column percentages reported.
<sup>2</sup>Pearson's Chi-squared test

Environmental exposures were each reported by about one in five participants: ETSE (19%, *n* = 506), ENAE (19%, *n* = 504), and ECSE (20%, *n* = 526). However, ETSE was higher in early (25%, *n* = 54) and middle adolescence (23%, *n* = 267), compared to late adolescence/young adulthood (14%, *n* = 185), whereas ENAE and ECSE were highest among those in late adolescence/young adulthood (22%, *n* = 285 and 26%, *n* = 342, respectively).

Among participants who experienced environmental exposures, daily proximity and household exposure were comparable across exposure types. Those reporting ETSE spent an average of 5.2 hours/day (*SD* = 6.80) near someone smoking cigarettes/combustible tobacco products and lived with 1.84 (*SD* = 4.82) household members who smoked; corresponding values were 4.22 hours/day (*SD* = 5.20) and 2.10 members (*SD* = 5.89) for ENAE, and 4.38 hours/day (*SD* = 5.64) and 2.36 members (*SD* = 6.31) for ECSE.

### Polyexposure and polyuse

Supplementary Table S6. presents combinations of substance use and environmental exposure. Substance use patterns differed significantly by age, χ²(14, *N* = 2,773) = 373.05, *p* < .001, Cramér’s *V* = .259. Most participants reported no current tobacco, ENDS, or cannabis use (58.9%, *n* = 1,634). Exclusive cannabis use was the most common use pattern (11.0%, *n* = 306), followed by dual ENDS-cannabis use (10.5%, *n* = 291), and then polyuse of cannabis, ENDS, and tobacco (9.7%, *n* = 270).

Environmental exposure combinations also varied by age, χ²(14, *N* = 2,619) = 160.01, *p* < .001, though the effect size was small (Cramér’s *V* = .175). Most respondents reported no exposure (63.5%). Exclusive ETSE was the most common pattern (7.6%, *n* = 199), peaking in middle adolescence (10.8%, *n* = 123) and lowest in late adolescence/young adulthood (4.5%, *n* = 58). Exclusive ECSE followed (6.9%, *n* = 181), peaking among late adolescents/young adults (11.0%, *n* = 140). Concurrent exposure to all three sources was the most common polyexposure pattern, reported by 4.8% (*n*=126) of the sample.

### Multiple group latent class analysis

Figure 2 presents the latent class item-response probabilities for the multiple group LCA with 4 classes. **Class 1: No Use/No Exposure (53.3%, *n*=1513)** was characterized by low probabilities of tobacco, ENDS, and cannabis use, as well as minimal exposure to cigarette, ENDS, and cannabis emissions. **Class 2: No Use/Polyexposure (10%, n=286)** reflected low probabilities of tobacco, ENDS, and cannabis use, but a high probability of ETSE, and moderate probabilities of ENAE and ECSE. **Class 3: Experimental Use/Low Exposure (22.3%, *n*=631)** demonstrated a low probability of tobacco use, moderate probabilities of experimental ENDS use and experimental cannabis use, and low probabilities of ETSE, ENAE, and ECSE. **Class 4: Polysubstance Use/Polyexposure (14.3%, *n*=407)** represented the highest risk profile, characterized by higher probabilities of current ENDS and cannabis use, lifetime tobacco use, and moderate probabilities of polyexposures.

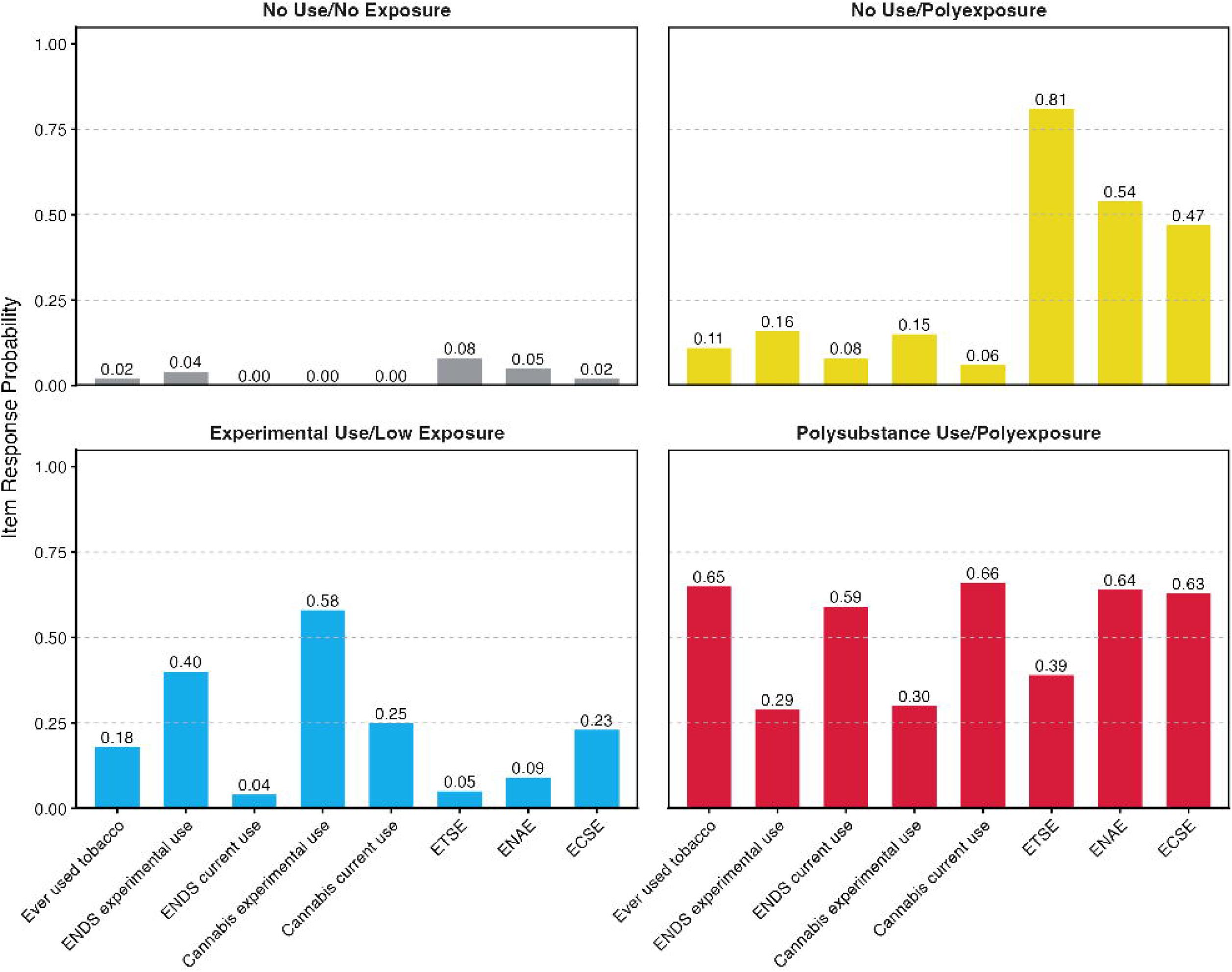

Figure 3 presents latent class membership prevalences across age groups. The No Use/No Exposure class was the most prevalent across all age groups (53%) and class membership was highest at 75% in early adolescence, then decreased to 67% in middle adolescence, and then 37% in late adolescence/young adulthood. The No Use/Polyexposure class included 10% of the sample and the estimated membership prevalence was highest at 22% in early adolescence, then 13% in middle adolescence, and 6% in late adolescence/young adulthood. The Experimental Use/Low Exposure Class was the second largest class including 22% of the sample and membership was lowest in early adolescence at 3%, to 11% in middle adolescence, and 36% in late adolescence/young adulthood. The Polysubstance Use/Polyexposure class included 14.3% of the sample and an estimated 9% of the middle adolescence group and 21% of the late adolescence/young adult group belonged to this class.

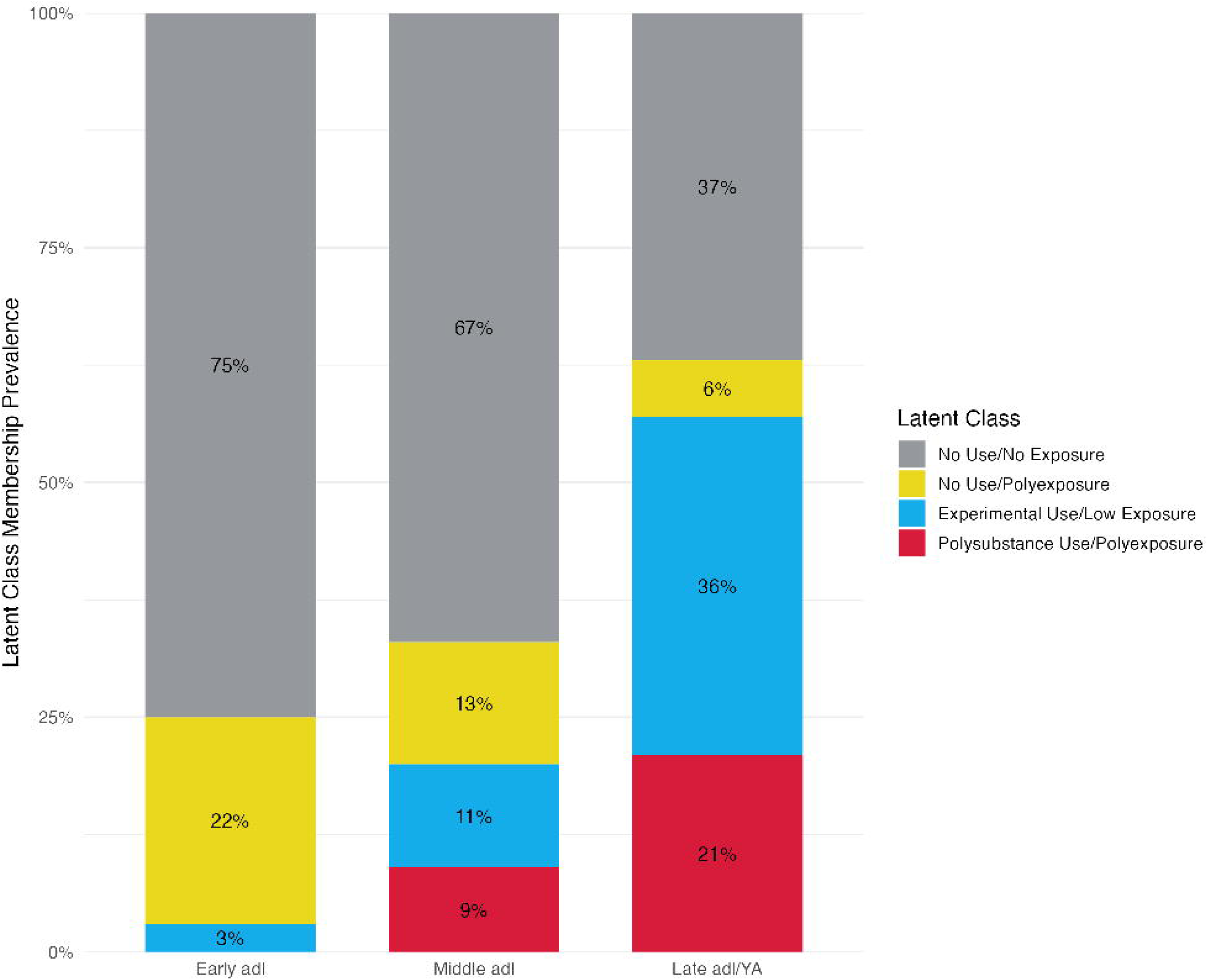

## DISCUSSION

We used a multiple group latent class analysis to identify patterns of cannabis, ENDS, and tobacco use, and environmental exposures to these substances, among adolescents and young adults enrolled in the Environmental influences on Child Health Outcomes (ECHO) study. Recent latent class studies of adolescent tobacco and cannabis use have shown increasingly complex patterns of polyuse,^23,24^ which our study extends by simultaneously characterizing substance use and environmental exposures. Four classes emerged: (1) No Use/No Exposure, (2) No Use/Polyexposure, (3) Experimental Use/Low Exposure, and (4) Polysubstance Use/Polyexposure. Class membership prevalence varied by stage of adolescence (early [11-13 years], middle [14-17 years], late adolescence/young adulthood [18-24 years]), with the No Use/Polyexposure class being most prevalent in early adolescence and the Polysubstance Use/Polyexposure class being most prevalent in late adolescence/young adulthood.

Among late adolescents and young adults, exclusive cannabis use was the most frequent pattern of substance use reported, followed by co-use of cannabis, ENDS, and tobacco products. This was reflected in their class membership prevalences, as 36% belonged to the Experimental Use/Low Exposure group and 21% belonged to the Polysubstance Use/Polyexposure group. These findings are consistent with national surveillance data showing increased use of cannabis and nicotine use among late adolescence and young adulthood.^25,26^ Notably, 26% also reported regular exposure to cannabis smoke, 22% reported ENAE, and 14% reported ETSE. Our findings suggest that environmental emissions exposure frequently co-occurs with polysubstance use among late adolescents and young adults.

Early adolescents had the highest-class membership to the No Use/Polyexposure class (22%), indicating that, despite not using substances themselves, over one-fourth were still exposed to environmental tobacco and cannabis emissions regularly. This finding is quite concerning, given the potential for increased health risks associated with chronic environmental exposures,^27,28^ and likely reflects exposure within the household from family members. Even if early adolescents are not using tobacco and cannabis products themselves, such exposure may reflect social modeling of substance use behaviors by parents and peers and place adolescents at risk for initiation later in adolescence.^29,30^ Further research with larger samples of early adolescents is warranted.

The source and composition of environmental exposures shifted across development. Early adolescents’ environmental exposures were predominantly tobacco-based and largely independent of their own behavior, whereas exposure among older adolescents was increasingly cannabis-based and more likely to co-occur with personal use. Although the cross-sectional design prevents us from determining whether exposure precedes, follows, or reciprocally influences use, our findings support a developmentally staged approach to adolescent prevention. For early adolescents, family- and household-level interventions may be most useful. Pediatric screening could assess environmental exposure in addition to personal use, while caregiver cessation support and family education could address the risks associated with tobacco, cannabis, and ENDS emissions.^32,33^ Encouraging parents to set household rules is another important target for prevention. Setting household rules banning indoor smoking and vaping is associated with greater perceived harm, lower cannabis and ENDS use, and reduced likelihood of tobacco initiation among adolescents.^11,27,35^ Most U.S. caregivers prohibit tobacco smoking in the home,^34^ whereas comparable protections for cannabis and ENDS use have lagged behind. In sum, using similar strategies for early and middle adolescents that have been effective for tobacco prevention shows promise for preventing cannabis and ENDS use and environmental exposures.

Among late adolescents and young adults, prevention may benefit from directly addressing cannabis-ENDS co-use and the social settings in which these behaviors occur. For instance, college campus-level tobacco-control policies have been associated with reductions in cigarette use and secondhand smoke exposure among college students, although they coincided with increased ENDS use.^36^ Evidence for campus cannabis policies is more limited, with preliminary evidence suggesting that sanctions following cannabis-policy violations may reduce subsequent use, but few studies have evaluated the population-level effects of campus-wide cannabis bans.^37^ Finally, continuing to develop and implement adolescent and young adult-specific treatment interventions that address polyuse and polyexposure (e.g., cognitive behavioral therapy for polyuse), and are easily accessible through technology, could lead to more fruitful prevention and treatment efforts.^38,39^

### Limitations and future directions

Several limitations should be considered while interpreting our findings. First, the cross-sectional design precludes causal inferences between substance use and environmental exposure patterns. The observed associations are compatible with several possibilities, including exposure preceding initiation, personal use generating subsequent exposure, or reciprocal processes in which exposure and use reinforce one another. Forthcoming ECHO data releases will permit longitudinal examination of the frequency, duration, and co-occurrence of use and exposure. Second, self-report measures were used which may introduce recall or reporting error. Future research should incorporate biometric data to confirm self-report measures. Third, environmental exposure levels and risks depend on factors such as ventilation, proximity, and product type (e.g., joints, blunts, or water pipes), which were not captured in this study.^14,40^ Identifying the most common locations and sources of environmental exposures is an important next step for prevention. Similarly, future work should examine correlates of class membership (housing type, SES, geography, family/peer use). Finally, while participants were drawn from 19 study sites across the US, the sample is not nationally representative. Replication in nationally representative samples will therefore be important for evaluating the generalizability of these developmental patterns.

### Conclusion

Our study demonstrates the importance of assessing tobacco, cannabis, and ENDS environmental exposures alongside adolescent substance use, and incorporating developmental context. Prevention targets should align with the stage of adolescence, with a focus on reducing household and caregiver-generated environmental exposures in early adolescence to addressing peer, social, and personal-use contexts in later adolescence and young adulthood. While further research is needed to better understand the risks associated with polyexposures, we should not hesitate to implement evidence-based prevention strategies that safeguard the health of today’s youth.

## Data Availability

Per the data use agreement, the authors do not have permission to release these data. Data requests can be submitted, learn more here: https://echochildren.org/data-availability/.

