## Supplementary Table 1 for "Latent Classes of Cannabis, ENDS, and Tobacco Use and Environmental Exposure Among Adolescents and Young Adults in the ECHO Cohort"

**Supplementary Table S1.** *Substance Use and Environmental Exposures Stratified by Survey Year.*

| **Variable** | **Overall**  N = 2,876  ¹ | **2018**  N = 92  ¹ | **2019**  N = 667  ¹ | **2020**  N = 948  ¹ | **2021**  N = 954  ¹ | **2022**  N = 214  ¹ |
| --- | --- | --- | --- | --- | --- | --- |
| **ENDS Frequency** |  |  |  |  |  |  |
| Never | 2,047 (74%) | 51 (86%) | 433 (67%) | 701 (75%) | 695 (74%) | 167 (79%) |
| Experimental | 454 (16%) | 4 (6.8%) | 140 (22%) | 144 (15%) | 145 (16%) | 21 (9.9%) |
| Current use | 282 (10%) | 4 (6.8%) | 73 (11%) | 87 (9.3%) | 94 (10%) | 24 (11%) |
| *Missing* | *93* | *33* | *21* | *16* | *20* | *2* |
| **Cannabis Frequency** |  |  |  |  |  |  |
| Never | 1,852 (66%) | 38 (42%) | 362 (55%) | 643 (69%) | 654 (70%) | 155 (74%) |
| Experimental | 530 (19%) | 30 (33%) | 176 (27%) | 154 (17%) | 145 (16%) | 25 (12%) |
| Current use | 436 (15%) | 23 (25%) | 116 (18%) | 135 (14%) | 132 (14%) | 29 (14%) |
| *Missing* | *58* | *1* | *13* | *16* | *23* | *5* |
| **Tobacco Frequency** |  |  |  |  |  |  |
| Never | 2,397 (85%) | 64 (70%) | 528 (80%) | 802 (86%) | 817 (87%) | 185 (87%) |
| Experimental | 339 (12%) | 22 (24%) | 102 (16%) | 104 (11%) | 89 (9.5%) | 22 (10%) |
| Current use | 97 (3.4%) | 6 (6.5%) | 26 (4.0%) | 28 (3.0%) | 31 (3.3%) | 6 (2.8%) |
| *Missing* | *43* | *0* | *11* | *14* | *17* | *1* |
| **ETSE** |  |  |  |  |  |  |
| No Exposure | 2,157 (81%) | 74 (85%) | 487 (84%) | 730 (82%) | 713 (79%) | 152 (75%) |
| Any Exposure | 506 (19%) | 13 (15%) | 95 (16%) | 160 (18%) | 187 (21%) | 51 (25%) |
| *Missing* | *213* | *5* | *85* | *58* | *54* | *11* |
| **ENAE** |  |  |  |  |  |  |
| No Exposure | 2,162 (81%) | 63 (72%) | 469 (80%) | 753 (85%) | 722 (80%) | 154 (76%) |
| Any Exposure | 504 (19%) | 25 (28%) | 120 (20%) | 136 (15%) | 175 (20%) | 48 (24%) |
| *Missing* | *210* | *4* | *78* | *59* | *57* | *12* |
| **ECSE** |  |  |  |  |  |  |
| No Exposure | 2,138 (80%) | 61 (73%) | 467 (79%) | 726 (82%) | 728 (81%) | 156 (78%) |
| Any Exposure | 526 (20%) | 23 (27%) | 122 (21%) | 160 (18%) | 175 (19%) | 45 (22%) |
| *Missing* | *212* | *8* | *78* | *62* | *51* | *13* |

***Note.*** ENDS= Electronic nicotine delivery system, ETSE=Environmental tobacco smoke exposure, ENAE=Environmental nicotine aerosols exposure, ECSE=Environmental cannabis smoke exposure.
**¹** n (%). Percentages were calculated excluding missing values unless otherwise indicated. “Any exposure” indicates self-reported environmental exposure. Missing indicates missing data.
