## Supplementary Table 2 for "Latent Classes of Cannabis, ENDS, and Tobacco Use and Environmental Exposure Among Adolescents and Young Adults in the ECHO Cohort"

**Supplementary Table S2.** *Substance Use and Environmental Exposures Stratified by Birth Year*

| **Variable** | **Overall**  N = 2,876  ¹ | **1994**  N = 1  ¹ | **1995**  N = 8  ¹ | **1996**  N = 66  ¹ | **1997**  N = 54  ¹ | **1998**  N = 17  ¹ | **1999**  N = 193  ¹ | **2000**  N = 314  ¹ | **2001**  N = 342  ¹ | **2002**  N = 374  ¹ | **2003**  N = 740  ¹ | **2004**  N = 330  ¹ | **2005**  N = 140  ¹ | **2006**  N = 60  ¹ | **2007**  N = 66  ¹ | **2008**  N = 91  ¹ | **2009**  N = 62  ¹ | **2010**  N = 15  ¹ | **2011**  N = 3  ¹ |
| --- | --- | --- | --- | --- | --- | --- | --- | --- | --- | --- | --- | --- | --- | --- | --- | --- | --- | --- | --- |
| **ENDS Frequency** |  |  |  |  |  |  |  |  |  |  |  |  |  |  |  |  |  |  |  |
| Never | 2,047 (74%) | 1 (100%) | 4 (57%) | 30 (73%) | 28 (74%) | 10 (59%) | 105 (58%) | 188 (62%) | 215 (64%) | 276 (74%) | 551 (75%) | 260 (79%) | 111 (82%) | 53 (93%) | 61 (92%) | 82 (91%) | 56 (95%) | 13 (93%) | 3 (100%) |
| Experimental | 454 (16%) | 0 (0%) | 2 (29%) | 5 (12%) | 7 (18%) | 6 (35%) | 51 (28%) | 63 (21%) | 80 (24%) | 59 (16%) | 110 (15%) | 37 (11%) | 19 (14%) | 3 (5.3%) | 3 (4.5%) | 5 (5.6%) | 3 (5.1%) | 1 (7.1%) | 0 (0%) |
| Current use | 282 (10%) | 0 (0%) | 1 (14%) | 6 (15%) | 3 (7.9%) | 1 (5.9%) | 25 (14%) | 54 (18%) | 40 (12%) | 37 (9.9%) | 72 (9.8%) | 31 (9.5%) | 6 (4.4%) | 1 (1.8%) | 2 (3.0%) | 3 (3.3%) | 0 (0%) | 0 (0%) | 0 (0%) |
| *Missing* | *93* | *0* | *1* | *25* | *16* | *0* | *12* | *9* | *7* | *2* | *7* | *2* | *4* | *3* | *0* | *1* | *3* | *1* | *0* |
| **Cannabis Frequency** |  |  |  |  |  |  |  |  |  |  |  |  |  |  |  |  |  |  |  |
| Never | 1,852 (66%) | 0 (0%) | 3 (38%) | 16 (25%) | 15 (28%) | 9 (53%) | 74 (41%) | 118 (39%) | 171 (51%) | 239 (64%) | 568 (77%) | 266 (82%) | 107 (78%) | 49 (86%) | 62 (94%) | 81 (91%) | 58 (98%) | 14 (100%) | 2 (100%) |
| Experimental | 530 (19%) | 0 (0%) | 1 (13%) | 26 (40%) | 27 (50%) | 5 (29%) | 57 (31%) | 102 (33%) | 85 (25%) | 79 (21%) | 84 (11%) | 28 (8.7%) | 19 (14%) | 6 (11%) | 3 (4.5%) | 7 (7.9%) | 1 (1.7%) | 0 (0%) | 0 (0%) |
| Current use | 436 (15%) | 1 (100%) | 4 (50%) | 23 (35%) | 12 (22%) | 3 (18%) | 50 (28%) | 85 (28%) | 79 (24%) | 53 (14%) | 82 (11%) | 29 (9.0%) | 11 (8.0%) | 2 (3.5%) | 1 (1.5%) | 1 (1.1%) | 0 (0%) | 0 (0%) | 0 (0%) |
| *Missing* | *58* | *0* | *0* | *1* | *0* | *0* | *12* | *9* | *7* | *3* | *6* | *7* | *3* | *3* | *0* | *2* | *3* | *1* | *1* |
| **Tobacco Frequency** |  |  |  |  |  |  |  |  |  |  |  |  |  |  |  |  |  |  |  |
| Never | 2,397 (85%) | 1 (100%) | 3 (38%) | 36 (55%) | 36 (67%) | 16 (94%) | 131 (73%) | 230 (75%) | 278 (83%) | 331 (89%) | 639 (87%) | 292 (89%) | 127 (91%) | 52 (91%) | 64 (97%) | 83 (91%) | 61 (98%) | 14 (93%) | 3 (100%) |
| Experimental | 339 (12%) | 0 (0%) | 4 (50%) | 25 (38%) | 13 (24%) | 1 (5.9%) | 39 (22%) | 60 (20%) | 49 (15%) | 30 (8.1%) | 74 (10%) | 24 (7.3%) | 7 (5.0%) | 4 (7.0%) | 1 (1.5%) | 6 (6.6%) | 1 (1.6%) | 1 (6.7%) | 0 (0%) |
| Current use | 97 (3.4%) | 0 (0%) | 1 (13%) | 5 (7.6%) | 5 (9.3%) | 0 (0%) | 10 (5.6%) | 15 (4.9%) | 7 (2.1%) | 11 (3.0%) | 22 (3.0%) | 12 (3.7%) | 5 (3.6%) | 1 (1.8%) | 1 (1.5%) | 2 (2.2%) | 0 (0%) | 0 (0%) | 0 (0%) |
| *Missing* | *43* | *0* | *0* | *0* | *0* | *0* | *13* | *9* | *8* | *2* | *5* | *2* | *1* | *3* | *0* | *0* | *0* | *0* | *0* |
| **ETSE** |  |  |  |  |  |  |  |  |  |  |  |  |  |  |  |  |  |  |  |
| No Exposure | 2,157 (81%) | 0 (0%) | 6 (86%) | 52 (81%) | 46 (85%) | 17 (100%) | 154 (89%) | 250 (90%) | 251 (86%) | 282 (82%) | 544 (76%) | 253 (80%) | 91 (75%) | 42 (78%) | 46 (72%) | 74 (85%) | 36 (63%) | 12 (86%) | 1 (33%) |
| Any Exposure | 506 (19%) | 0 (0%) | 1 (14%) | 12 (19%) | 8 (15%) | 0 (0%) | 20 (11%) | 29 (10%) | 41 (14%) | 60 (18%) | 172 (24%) | 64 (20%) | 31 (25%) | 12 (22%) | 18 (28%) | 13 (15%) | 21 (37%) | 2 (14%) | 2 (67%) |
| *Missing* | *213* | *1* | *1* | *2* | *0* | *0* | *19* | *35* | *50* | *32* | *24* | *13* | *18* | *6* | *2* | *4* | *5* | *1* | *0* |
| **ENAE** |  |  |  |  |  |  |  |  |  |  |  |  |  |  |  |  |  |  |  |
| No Exposure | 2,162 (81%) | 0 (0%) | 5 (63%) | 43 (68%) | 41 (76%) | 12 (71%) | 138 (78%) | 215 (76%) | 230 (78%) | 282 (83%) | 595 (83%) | 263 (84%) | 107 (88%) | 46 (87%) | 54 (83%) | 76 (87%) | 42 (75%) | 12 (86%) | 1 (33%) |
| Any Exposure | 504 (19%) | 0 (0%) | 3 (38%) | 20 (32%) | 13 (24%) | 5 (29%) | 39 (22%) | 67 (24%) | 63 (22%) | 58 (17%) | 124 (17%) | 51 (16%) | 14 (12%) | 7 (13%) | 11 (17%) | 11 (13%) | 14 (25%) | 2 (14%) | 2 (67%) |
| *Missing* | *210* | *1* | *0* | *3* | *0* | *0* | *16* | *32* | *49* | *34* | *21* | *16* | *19* | *7* | *1* | *4* | *6* | *1* | *0* |
| **ECSE** |  |  |  |  |  |  |  |  |  |  |  |  |  |  |  |  |  |  |  |
| No Exposure | 2,138 (80%) | 0 (0%) | 4 (57%) | 41 (66%) | 39 (72%) | 13 (76%) | 121 (69%) | 200 (70%) | 221 (76%) | 277 (81%) | 615 (85%) | 285 (91%) | 92 (75%) | 44 (81%) | 51 (80%) | 74 (90%) | 47 (84%) | 12 (92%) | 2 (100%) |
| Any Exposure | 526 (20%) | 1 (100%) | 3 (43%) | 21 (34%) | 15 (28%) | 4 (24%) | 55 (31%) | 84 (30%) | 71 (24%) | 65 (19%) | 107 (15%) | 29 (9.2%) | 30 (25%) | 10 (19%) | 13 (20%) | 8 (9.8%) | 9 (16%) | 1 (7.7%) | 0 (0%) |
| *Missing* | *212* | *0* | *1* | *4* | *0* | *0* | *17* | *30* | *50* | *32* | *18* | *16* | *18* | *6* | *2* | *9* | *6* | *2* | *1* |
| Note. ENDS= Electronic nicotine delivery system, ETSE=Environmental tobacco smoke exposure, ENAE=Environmental nicotine aerosols exposure, ECSE=Environmental cannabis smoke exposure. | | | | | | | | | | | | | | | | | | | |

¹ n (%)
