## Supplementary Table 3 for "Latent Classes of Cannabis, ENDS, and Tobacco Use and Environmental Exposure Among Adolescents and Young Adults in the ECHO Cohort"

**Supplementary Table S3.** *Substance Use and Environmental Exposures Stratified by Cohort.*

|  | **Overall**, N = 2,876^1^ | **AAC01**, N = 133^1^ | **AAD01**, N = 28^1^ | **AAE02**, N = 50^1^ | **AAJ01**, N = 151^1^ | **AAL01**, N = 305^1^ | **AAP02**, N = 129^1^ | **AAT01**, N = 339^1^ | **AAU01**, N = 599^1^ | **AAX03**, N = 64^1^ | **AAX04**, N = 94^1^ | **AAX05**, N = 167^1^ | **AAZ01**, N = 103^1^ | **ABA01**, N = 8^1^ | **ABA02**, N = 20^1^ | **ABA03**, N = 12^1^ | **ABA05**, N = 16^1^ | **ACA01**, N = 41^1^ | **AEA01**, N = 611^1^ | **AJA01**, N = 6^1^ |
| --- | --- | --- | --- | --- | --- | --- | --- | --- | --- | --- | --- | --- | --- | --- | --- | --- | --- | --- | --- | --- |
| **ENDS Frequency** |  |  |  |  |  |  |  |  |  |  |  |  |  |  |  |  |  |  |  |  |
| Never | 2,047 (74%) | 115 (86%) | 25 (89%) | 41 (85%) | 100 (67%) | 230 (78%) | 63 (72%) | 283 (84%) | 345 (61%) | 33 (52%) | 76 (81%) | 103 (62%) | 98 (96%) | 7 (88%) | 18 (95%) | 11 (92%) | 14 (88%) | 36 (90%) | 443 (73%) | 6 (100%) |
| Experimental | 454 (16%) | 16 (12%) | 2 (7.1%) | 6 (13%) | 28 (19%) | 45 (15%) | 14 (16%) | 34 (10%) | 142 (25%) | 22 (34%) | 13 (14%) | 39 (23%) | 2 (2.0%) | 1 (13%) | 1 (5.3%) | 0 (0%) | 1 (6.3%) | 3 (7.5%) | 85 (14%) | 0 (0%) |
| Current use | 282 (10%) | 2 (1.5%) | 1 (3.6%) | 1 (2.1%) | 22 (15%) | 21 (7.1%) | 10 (11%) | 20 (5.9%) | 83 (15%) | 9 (14%) | 5 (5.3%) | 25 (15%) | 2 (2.0%) | 0 (0%) | 0 (0%) | 1 (8.3%) | 1 (6.3%) | 1 (2.5%) | 78 (13%) | 0 (0%) |
| Missing | 93 | 0 | 0 | 2 | 1 | 9 | 42 | 2 | 29 | 0 | 0 | 0 | 1 | 0 | 1 | 0 | 0 | 1 | 5 | 0 |
| **Cannabis Frequency** |  |  |  |  |  |  |  |  |  |  |  |  |  |  |  |  |  |  |  |  |
| Never | 1,852 (66%) | 107 (80%) | 26 (93%) | 42 (91%) | 73 (49%) | 246 (83%) | 34 (27%) | 273 (81%) | 219 (38%) | 27 (42%) | 60 (64%) | 102 (61%) | 97 (96%) | 7 (88%) | 20 (100%) | 12 (100%) | 15 (94%) | 35 (88%) | 451 (75%) | 6 (100%) |
| Experimental | 530 (19%) | 18 (14%) | 1 (3.6%) | 4 (8.7%) | 30 (20%) | 31 (11%) | 54 (42%) | 37 (11%) | 197 (35%) | 19 (30%) | 18 (19%) | 34 (20%) | 2 (2.0%) | 1 (13%) | 0 (0%) | 0 (0%) | 0 (0%) | 5 (13%) | 79 (13%) | 0 (0%) |
| Current use | 436 (15%) | 8 (6.0%) | 1 (3.6%) | 0 (0%) | 47 (31%) | 18 (6.1%) | 40 (31%) | 26 (7.7%) | 154 (27%) | 18 (28%) | 16 (17%) | 31 (19%) | 2 (2.0%) | 0 (0%) | 0 (0%) | 0 (0%) | 1 (6.3%) | 0 (0%) | 74 (12%) | 0 (0%) |
| Missing | 58 | 0 | 0 | 4 | 1 | 10 | 1 | 3 | 29 | 0 | 0 | 0 | 2 | 0 | 0 | 0 | 0 | 1 | 7 | 0 |
| **Tobacco Frequency** |  |  |  |  |  |  |  |  |  |  |  |  |  |  |  |  |  |  |  |  |
| Never | 2,397 (85%) | 125 (94%) | 26 (93%) | 48 (96%) | 134 (89%) | 274 (91%) | 76 (59%) | 314 (93%) | 435 (77%) | 49 (77%) | 82 (87%) | 129 (77%) | 100 (97%) | 8 (100%) | 20 (100%) | 12 (100%) | 14 (88%) | 34 (83%) | 511 (84%) | 6 (100%) |
| Experimental | 339 (12%) | 7 (5.3%) | 1 (3.6%) | 2 (4.0%) | 11 (7.3%) | 24 (8.0%) | 42 (33%) | 19 (5.6%) | 114 (20%) | 10 (16%) | 7 (7.4%) | 32 (19%) | 1 (1.0%) | 0 (0%) | 0 (0%) | 0 (0%) | 1 (6.3%) | 6 (15%) | 62 (10%) | 0 (0%) |
| Current use | 97 (3.4%) | 1 (0.8%) | 1 (3.6%) | 0 (0%) | 6 (4.0%) | 3 (1.0%) | 11 (8.5%) | 4 (1.2%) | 19 (3.3%) | 5 (7.8%) | 5 (5.3%) | 6 (3.6%) | 2 (1.9%) | 0 (0%) | 0 (0%) | 0 (0%) | 1 (6.3%) | 1 (2.4%) | 32 (5.3%) | 0 (0%) |
| Missing | 43 | 0 | 0 | 0 | 0 | 4 | 0 | 2 | 31 | 0 | 0 | 0 | 0 | 0 | 0 | 0 | 0 | 0 | 6 | 0 |
| **ETSE** |  |  |  |  |  |  |  |  |  |  |  |  |  |  |  |  |  |  |  |  |
| No Exposure | 2,157 (81%) | 119 (92%) | 27 (96%) | 33 (66%) | 118 (84%) | 229 (87%) | 104 (83%) | 258 (79%) | 447 (91%) | 55 (90%) | 56 (61%) | 144 (87%) | 65 (67%) | 8 (100%) | 16 (84%) | 12 (100%) | 15 (94%) | 32 (86%) | 414 (69%) | 5 (83%) |
| Any Exposure | 506 (19%) | 11 (8.5%) | 1 (3.6%) | 17 (34%) | 22 (16%) | 34 (13%) | 21 (17%) | 68 (21%) | 43 (8.8%) | 6 (9.8%) | 36 (39%) | 21 (13%) | 32 (33%) | 0 (0%) | 3 (16%) | 0 (0%) | 1 (6.3%) | 5 (14%) | 184 (31%) | 1 (17%) |
| Missing | 213 | 3 | 0 | 0 | 11 | 42 | 4 | 13 | 109 | 3 | 2 | 2 | 6 | 0 | 1 | 0 | 0 | 4 | 13 | 0 |
| **ENAE** |  |  |  |  |  |  |  |  |  |  |  |  |  |  |  |  |  |  |  |  |
| No Exposure | 2,162 (81%) | 126 (95%) | 27 (96%) | 34 (68%) | 113 (80%) | 222 (86%) | 89 (71%) | 287 (88%) | 384 (78%) | 48 (75%) | 78 (84%) | 118 (72%) | 75 (77%) | 8 (100%) | 18 (95%) | 11 (100%) | 13 (81%) | 35 (95%) | 471 (79%) | 5 (83%) |
| Any Exposure | 504 (19%) | 6 (4.5%) | 1 (3.6%) | 16 (32%) | 28 (20%) | 37 (14%) | 36 (29%) | 41 (13%) | 107 (22%) | 16 (25%) | 15 (16%) | 46 (28%) | 23 (23%) | 0 (0%) | 1 (5.3%) | 0 (0%) | 3 (19%) | 2 (5.4%) | 125 (21%) | 1 (17%) |
| Missing | 210 | 1 | 0 | 0 | 10 | 46 | 4 | 11 | 108 | 0 | 1 | 3 | 5 | 0 | 1 | 1 | 0 | 4 | 15 | 0 |
| **ECSE** |  |  |  |  |  |  |  |  |  |  |  |  |  |  |  |  |  |  |  |  |
| No Exposure | 2,138 (80%) | 116 (87%) | 28 (100%) | 38 (81%) | 90 (62%) | 232 (90%) | 84 (68%) | 290 (88%) | 354 (72%) | 46 (72%) | 54 (57%) | 128 (79%) | 79 (81%) | 8 (100%) | 18 (95%) | 12 (100%) | 16 (100%) | 29 (94%) | 511 (85%) | 5 (83%) |
| Any Exposure | 526 (20%) | 17 (13%) | 0 (0%) | 9 (19%) | 55 (38%) | 26 (10%) | 40 (32%) | 40 (12%) | 135 (28%) | 18 (28%) | 40 (43%) | 35 (21%) | 19 (19%) | 0 (0%) | 1 (5.3%) | 0 (0%) | 0 (0%) | 2 (6.5%) | 88 (15%) | 1 (17%) |
| Missing | 212 | 0 | 0 | 3 | 6 | 47 | 5 | 9 | 110 | 0 | 0 | 4 | 5 | 0 | 1 | 0 | 0 | 10 | 12 | 0 |
| *^Note.^* ^ENDS= Electronic nicotine delivery system, ETSE=Environmental tobacco smoke exposure, ENAE=Environmental nicotine aerosols exposure, ECSE=Environmental cannabis smoke exposure.^ **^¹^** ^n (%). Percentages were calculated excluding missing values unless otherwise indicated. “Any exposure” indicates self-reported environmental exposure. Missing indicates missing data.^ | | | | | | | | | | | | | | | | | | | | |
