## Supplementary Table 4 for "Latent Classes of Cannabis, ENDS, and Tobacco Use and Environmental Exposure Among Adolescents and Young Adults in the ECHO Cohort"

**Supplementary Table S4**

*Fit Indices for Multiple-Group Latent Class Analysis Models With 2-6 Classes*

| *K* | *log L* | *AIC* | *BIC* | *G²* | *df* | *Entropy* |
| --- | --- | --- | --- | --- | --- | --- |
| 2 | -8,415.51 | 16,869.02 | 16,982.08 | 1,480.61 | 412 | .804 |
| 3 | -8,222.85 | 16,505.69 | 16,684.21 | 1,095.29 | 401 | .808 |
| **4** | **-8,059.00** | **16,200.00** | **16,443.97** | **767.60** | **390** | **.777** |
| 5 | -8,001.06 | 16,106.11 | 16,415.54 | 651.71 | 379 | .791 |
| 6 | -7,972.60 | 16,071.30 | 16,446.20 | 584.22 | 368 | .733 |

*Note.* K = number of latent classes; *log L* = log-likelihood; AIC = Akaike Information Criterion; BIC = Bayesian Information Criterion; G² = likelihood-ratio goodness-of-fit statistic; df = degrees of freedom. All models estimated with measurement invariance across age group and non-invariant class proportions.
