## Supplementary Table 5 for "Latent Classes of Cannabis, ENDS, and Tobacco Use and Environmental Exposure Among Adolescents and Young Adults in the ECHO Cohort"

**Supplementary Table S5**

*Test of Measurement Invariance Across Age Groups.*

| Model | Parameters | Log-likelihood | AIC | BIC | Entropy |
| --- | --- | --- | --- | --- | --- |
| Invariant | 41 | -8,059.0 | 16,200.0 | 16,444.0 | .78 |
| Non-invariant | 105 | -7,956.2 | 16,122.5 | 16,747.3 | .78 |

*Note.* The likelihood-ratio test indicated statistically significant non-invariance (ΔG² = 205.5, Δdf = 64, P < .001); however, this result is anticipated due to the sample size. The BIC favored the parsimonious invariant model (16,444 vs. 16,747), and the entropy (0.78) was the same for both models, indicating that freeing the 64 additional parameters did not improve classification. Therefore, the invariant model was retained.
