## Supplementary Table 6 for "Latent Classes of Cannabis, ENDS, and Tobacco Use and Environmental Exposure Among Adolescents and Young Adults in the ECHO Cohort"

**Supplementary Table S6.** *Combinations of Polyuse and Polyexposure Stratified by Age Group.*

| **Substance Use** |  |  |  |  |  |  |
| --- | --- | --- | --- | --- | --- | --- |
|  | **Overall**  **(*N* = 2,773)** | **Early Adolescence**  **(n = 218)** | **Middle Adolescence**  **(n = 1,228)** | **Late Adolescence/Young Adulthood**  **(n = 1,327)** | **p-value** | **Adjusted p-value** |
| None | 1634 (58.9%) | 196 (89.9%) | 861 (70.1) | 577 (43.5) | < .001 | < .001 |
| Exclusively tobacco | 36 (1.3) | 2 (0.9) | 23 (1.9) | 11 (0.8) | .056 | .168 |
| Exclusively ENDS | 135 (4.9%) | 7 (3.2%) | 71 (5.8%) | 57 (4.3%) | .108 | .217 |
| Exclusively cannabis | 306 (11%) | 1 (0.5%) | 71 (5.8%) | 234 (17.6%) | < .001 | < .001 |
| Tobacco + ENDS | 39 (1.4%) | 3 (1.4%) | 23 (1.9%) | 13 (1.0%) | .157 | .217 |
| Tobacco + Cannabis | 62 (2.2%) | 5 (2.3%) | 13 (1.1%) | 44 (3.3%) | < .001 | .001 |
| ENDS + Cannabis | 291 (10.5%) | 3 (1.4%) | 98 (8.0%) | 190 (14.3%) | < .001 | < .001 |
| Tobacco + ENDS + Cannabis | 270 (9.7%) | 1 (0.5) | 68 (5.5%) | 201 (15.1%) | < .001 | < .001 |
| *Note.* Cell values are *n* (column %), computed among respondents with complete use data (*N* = 2,773). Combinations are mutually exclusive and reflect any past use of the indicated products. The omnibus association with age group was statistically significant, *χ*²(14, *N* = 2,773) = 373.05, *p* < .001, Cramér’s *V* = .259. Per-row *p* values are from category-versus-rest tests across age group (2 × 3; Pearson *χ*², or Fisher’s exact where any expected count < 5); Holm’s method was used p-value was calculated to adjust for multiple testing across the eight combinations. ENDS=Electronic nicotine delivery systems. | | | | | | |
| **Environmental Exposures** | |  |  |  |  |  |
|  | **Overall**  ***(N* = 2,619)** | **Early Adolescence**  **(n = 205)** | **Middle Adolescence (n = 1,139)** | **Late Adolescence/Young Adulthood**  **(n = 1,275)** | **p-value** | **Adjusted p-value** |
| None | 1664 (63.5%) | 139 (67.8%) | 759 (66.6%) | 766 (60.1%) | .002 | .008 |
| Exclusively ETSE | 199 (7.6%) | 18 (8.8%) | 123 (10.8%) | 58 (4.5%) | < .001 | < .001 |
| Exclusively ENAE | 161 (6.1%) | 7 (3.4%) | 61 (5.4%) | 93 (7.3%) | .033 | .100 |
| Exclusively ECSE | 181 (6.9%) | 4 (2.0%) | 37 (3.2%) | 140 (11.0%) | < .001 | < .001 |
| ETSE + ENAE | 86 (3.3%) | 13 (6.3%) | 43 (3.8%) | 30 (2.4%) | .006 | .022 |
| ETSE + ECSE | 84 (3.2%) | 8 (3.9%) | 42 (3.7%) | 34 (2.7%) | .306 | .613 |
| ENAE + ECSE | 118 (4.5%) | 2 (1.0%) | 23 (2.0%) | 93 (7.3%) | < .001 | < .001 |
| ETSE + ENAE + ECSE | 126 (4.8%) | 14 (6.8%) | 51 (4.5%) | 61 (4.8%) | .350 | .613 |
| *Note.* Cell values are *n* (column %), computed among respondents with complete exposure data (*N* = 2,619). Combinations are mutually exclusive. The omnibus association with age group was statistically significant, *χ*²(14, *N* = 2,619) = 160.01, *p* < .001, Cramér’s *V* = .175. Per-row *p* values are from category-versus-rest tests across age group (2 × 3 Pearson *χ*²); Holm’s method was used to adjust for multiple tests.  ETSE = environmental tobacco smoke exposure; ENAE = environmental nicotine aerosol exposure; ECSE = environmental cannabis smoke exposure. | | | | | | |
